# The Impact of Operative Video Review versus Annotation on Training in Endoscopic Pituitary Surgery: A Preclinical Randomised Controlled Study

**DOI:** 10.1101/2024.11.01.24315810

**Authors:** Emalee J. Burrows, Joachim Starup-Hansen, Danyal Z. Khan, Chan Hee Koh, Maryam Hussain, James Booker, Danail Stoyanov, Hani J. Marcus

**Author notes:** Corresponding author: Dr Emalee Burrows, MRes Address: Queens Square, London, WC1N3BG, UK.

## Abstract

**Objective:** This study evaluated the effect of active operative video annotation on surgical education, specifically focusing on implementability, knowledge acquisition, skill development, and confidence.

**Background:** Resident duty hour restrictions necessitate more efficient surgical training, as steep learning curves in many procedures may result in residents completing training without gaining enough experience to perform them safely. Video annotation of operative videos, involving labeling of instruments and steps, might offer a secure and risk-free environment to improve surgical learning.

**Methods:** A preclinical randomized controlled trial was conducted with novice neurosurgeons from multiple centres. Participants were assigned to annotate real-life operative videos or to the control group, who performed passive video review. At baseline and then following either video annotation or video review, both groups completed a simulated pituitary adenoma resection on a validated high-fidelity physical simulator and were assessed using knowledge quizzes, a modified Global Rating Scale (mGRS), and confidence surveys. Participants also completed an implementability questionnaire.

**Results:** Fourteen participants completed the study. Psychometric surveys indicated 100% agreement on feasibility, acceptability, and appropriateness in the annotation group, significantly higher than the review group (p<0.001). Procedural knowledge score changes were significantly higher in the annotation group compared to the video review group (1.71, 95% CI: 0.19-3.41, p= 0.0479). The annotation group also significantly improved their operative performance from baseline, with mean mGRS increasing by 5.14 (95% CI: 2.36-7.93, p=0.004) versus 2.57 (95% CI: -1.30-6.44) (p=0.16) in the video review group. Confidence improved significantly in both groups (<0.05), with no significant difference between groups.

**Conclusions:** Active video annotation is a feasible and acceptable tool for enhancing surgical education. It led to a higher change in knowledge score compared to passive video review and also improved skills and confidence from baseline, suggesting its suitability for integration into surgical training programs. Its impact, however, on real-world surgical performance and patient outcomes requires further study.

## 1. Introduction

There exists a steep learning curve in surgical practice (1), which requires the input of thousands of hours of operative training to reach a stable plateau that achieves optimal patient outcomes (2). The COVID-19 pandemic (3, 4), recent changes to the Intercollegiate Surgical Curriculum Programme (ISCP) (5, 6), work-hour restrictions (7), and the development of increasingly complex operative techniques, such as robotic surgery (8), are all placing additional and increasing pressure on surgeons to train and maintain skills. This is also greater pressure on the healthcare system to find resource-effective methods to maintain high standards of patient care and surgical delivery. In light of these recent changes, the exposure of trainees to surgical operations and teaching opportunities is becoming increasingly limited (9).

There is evidence that deliberate practice of surgical technique and high-quality training outside of the operating theatre can impact the learning curve rates (10). This may also be applicable to experts, where deliberate practice through video or simulation training may be able to increase skills beyond the plateau obtained from operative experience and innate skill alone (11, 12). The review of operative videos for education is reported widely in the literature (13). However, most interactions with operative videos involve a one-way digestion of the content, with no interaction on the part of the learner. This may limit learning gains from a given operative video. The annotation of operative videos, which involves learners identifying key components of an operation, such as its anatomy, instruments, and steps, may offer an opportunity to more effectively learn in this setting. Specifically, such an intervention may offer a low-cost platform with the ability to review and interact with a display of the critical operative events at the user’s own pace (14).

This study proposes that the annotation of operative videos may provide a practical solution for enhancing surgical training. By offering a low-cost, interactive platform for reviewing and revising key operative steps, it aims to improve theoretical knowledge, practical skills, and confidence. We also aimed to evaluate whether annotating intraoperative surgical videos is feasible, acceptable, and appropriate for surgical training.

## 2. Methods

The study was a parallel randomised controlled trial to determine the effect of operative video annotation on surgical training. Ethics approval was obtained from the University College London Research Ethics Committee for studies on simulation (26117/002). The study was reported according to the Consolidated Standards of Reporting Trials (CONSORT) guidelines (15) and the accompanying simulation-based research extension (16).

A knowledge quiz, modified Global Rating Scale (mGRS) measurements, and a confidence survey were completed at baseline and after a 1-week period. During this time the intervention group was instructed to annotate videos, while the control group reviewed the videos. ChatGPT 4o was used to assist with wording and proofreading, with all output screened before publication.

### Participants

Novice surgeons were invited to participate through convenience sampling. Novice surgeons were defined as having obtained a medical degree and some surgical experience but not having completed a pituitary resection as the primary surgeon. The study was conducted at the National Hospital of Neurology and Neurosurgery, with data collected from June to July 2024. Demographic data, including age, biological sex, dominant hand, training grade, and operative experience, were collected.

### The annotation platform

The TouchSurgery^™^ enterprise application is a novel AI-powered surgical platform which has been designed to analyse surgical videos. The platform allows users to timestamp key procedural steps or errors using custom video tags (Figure 1). For this study the custom video tags focused on applying the Delphi consensus derived pituitary operative workflow analysis to the videos (17). Participants were required to annotate six videos using the workflow framework, including 40 steps and instruments. Anatomy annotation was not included due to platform constraints. Both groups were provided with the workflow in text form (17) and an instructional video on pituitary adenoma surgery.

**Figure 1.**
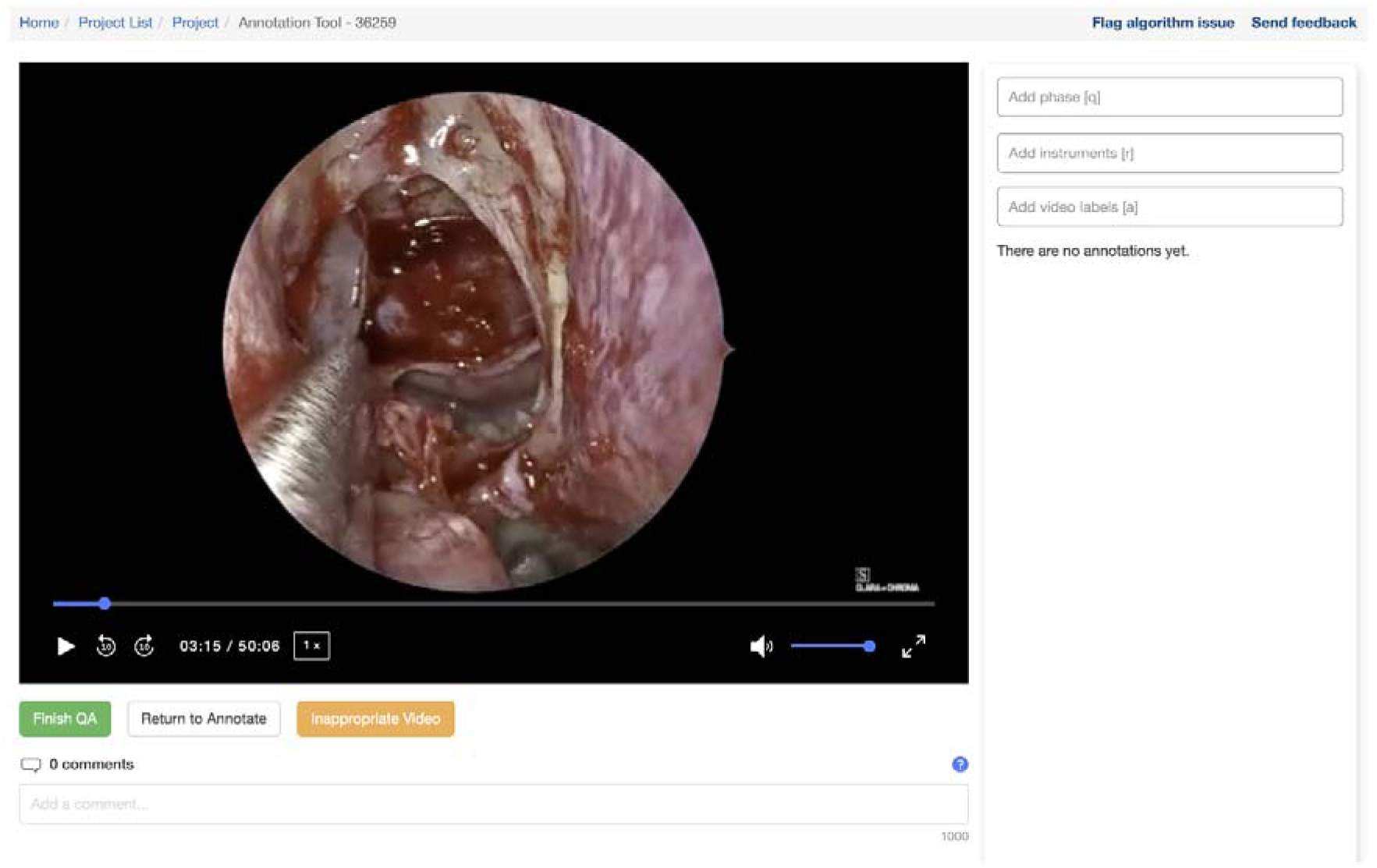
Annotation platform. The TouchSurgery^™^ enterprise interface with options for the participant to annotate the video via a drop-down box.

### Surgical task

Participants were required to perform the sellar phase of a simulated pituitary macroadenoma removal using a model. Operations were performed using a trans-sphenoidal box model. A sphenoidotomy had been pre-drilled prior to the assessment. Participants were allowed one attempt at the task, and no time limit was imposed. The entire endoscopic pituitary adenoma resection procedure was recorded through the endoscope.

### Outcomes: Feasibility, acceptability and appropriateness

The feasibility, acceptability, and appropriateness of both video annotation and passive video review were assessed though a validated psychometric assessment (18). It includes 12 questions rated on a Likert scale from 1=strongly disagree to 5=strongly agree. Feasibility assesses whether the new treatment or innovation can realistically be implemented in a given setting. Acceptability is a personal and incorporates a person’s expectations, needs and preferences. The appropriateness places the task in the technical social context, whether it achieves a given task and fits in with societal norms.

### Outcomes: knowledge acquisition

All participants completed knowledge tests assessing the domains of anatomical and procedural knowledge. A multiple-choice questionnaire assessed procedural knowledge and was adapted from a pituitary workflow framework (17). Here, participants watched a short video clip then was asked to identify the step and the associated instrument. Anatomical knowledge was tested using a previously developed anatomy outlining tool on an image of the sella, focussing on critical anatomical structures such as the carotid arteries and optic nerves (19).

### Outcomes: skill acquisition

Unedited videos of participants performing the sellar phase of the surgery were marked by two independent blinded assessors. Any discrepancies of more than two marks in a section were resolved by a third reviewer. A previously validated mGRS (20) was used.

### Outcomes: confidence

All participants completed surveys adapted from Bhattacharyya, Davidson (21) on self-reported confidence. Domains included confidence in the ability to perform a pituitary adenoma resection, identify anatomy, understand steps, make decisions, and identify errors. These were assessed using a five-point Likert scale, where 1=strongly disagree and 5=strongly agree.

### Randomisation and Statistical analysis

A priori power calculation was performed using raw data from a previous transsphenoidal pituitary adenoma surgery simulation conducted within our research group (22). The minimum clinically important effect was based on prior literature and expert consensus, with a conservative estimate of 25% (23). A power (1-β) of 80% and two-sided significance level (α) of 0.05 were used. This resulted in a required sample size of n=7 in each group. Participants were block-randomized into two equal groups (n=7) using the random number generator function in Microsoft Excel, with allocation concealment.

Data were evaluated for normality, and statistical methods were adjusted accordingly. All results were analysed on an intention-to-treat basis. Two-sided paired t-tests or Wilcoxon signed-rank tests were performed as appropriate to assess differences from baseline, and an unpaired t-test or Wilcoxon rank-sum test was used to assess differences between groups. Demographic characteristics were reported as percentages for categorical variables, with the remainder represented as median and interquartile range (IQR). The psychometric assessment was reported as percentage agreement, with a test of proportions. The odds for higher ratings were also calculated using an ordered logistic regression model. Analysis of confidence data was performed using a numerical rank (1 = Strongly Disagree, 2 = Disagree, 3 = Neither Agree nor Disagree, 4 = Agree, 5 = Strongly Agree) and calculating the median (IQR) for each statement.

## 3. Results

### Demographics

A total of 14 participants completed the study, with 7 in the intervention group and 7 in the control group (Figure 2). Four participants did not complete the baseline assessment. The demographic characteristics of participants are outlined in Table 1.

**Figure 2.**
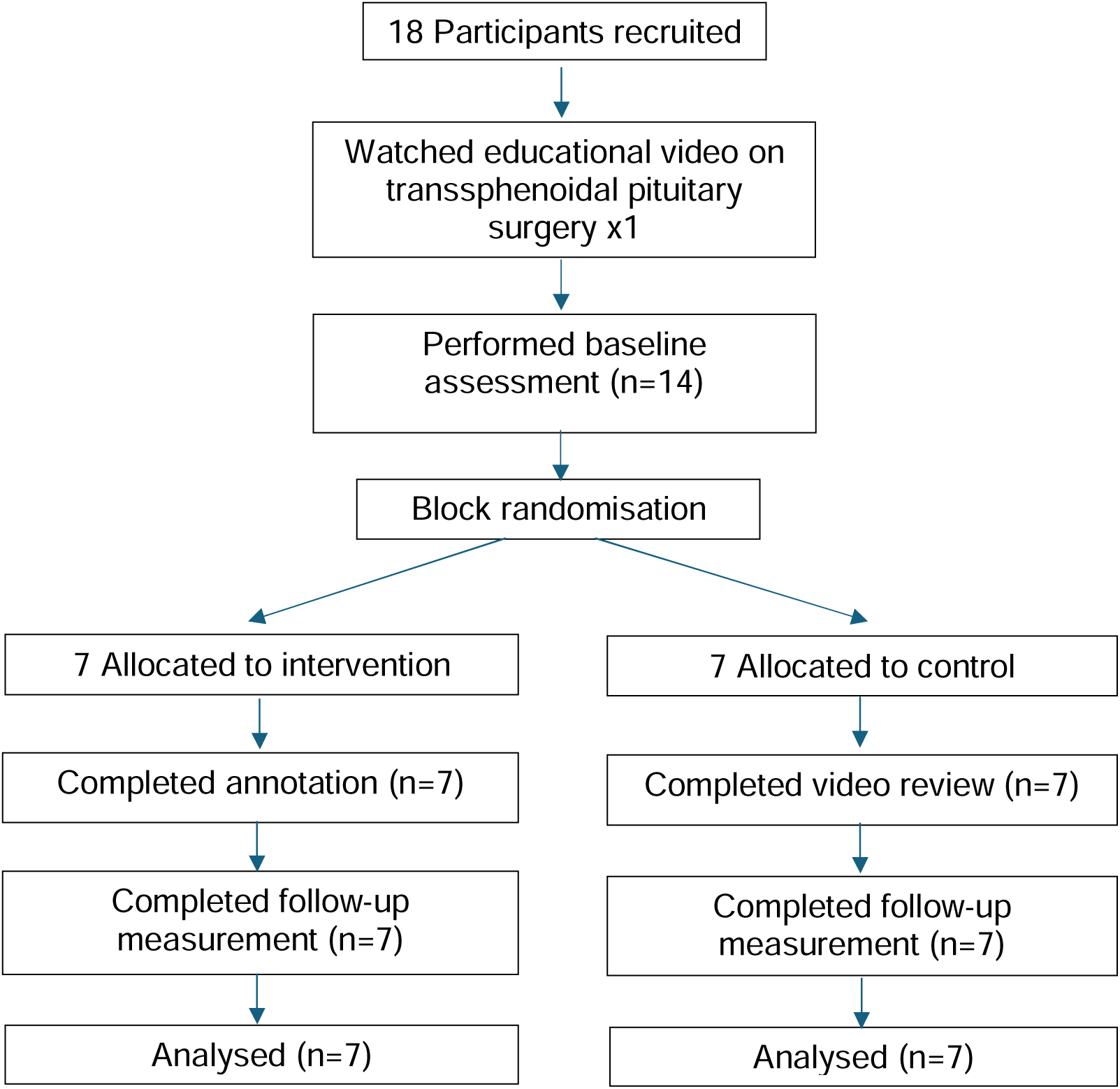
Flow chart of the study.

**Table 1.** Demographic Characteristics of Included Study Participants.

| Characteristic | Video annotation | Video review | p-value | Overall |
| --- | --- | --- | --- | --- |

|  | group | group |  |  |
| --- | --- | --- | --- | --- |
| **Age, median (IQR)** | 28 (26-31) | 28 (26-29) | 0.642 | 28 (26-29) |
| **Biological sex (%)** |  |  | 1.00 |  |
| -Female | 1 (14.2%) | 2 (28.6%) |  | 3 (21.4%) |
| -Male | 6 (85.7%) | 5 (71.4%) |  | 11 (78.6%) |
| **Training level (%)** |  |  | 0.356 |  |
| -Foundation Doctor | 2 (28.6%) | 0 (0%) |  | 2 (14.3%) |
| -SHO | 2 (28.6%) | 3 (42.9%) |  | 5 (35.7%) |
| -ST1-4 | 3 (42.9%) | 3 (42.9%) |  | 6 (42.9%) |
| -ST 5-8 | 0 (0%) | 1 (14.3%) |  | 1 (7.1%) |
| **Handedness (%)** |  |  | 1.0 |  |
| -Left | 0 (0%) | 0 (0%) |  | 0 (0%) |
| -Right | 8 (100%) | 8 (100%) |  | 16 (100%) |
| -Ambidextrous | 0 (0%) | 0 (0%) |  | 0 (0%) |
| **Transsphenoidal**<br>**Surgeries, median**<br>**(IQR)** |  |  |  |  |
| -number observed | 3.0 (3.0-5.0) | 4.0 (3.0-5.0) | 0.752 | 4 (3.0-5.0) |
| -number assisted | 3.0 (0.0-5.0) | 1.0 (0.0-3.0) | 0.671 | 1.5 (0.0-5.0) |

### Implementability Outcomes: Feasibility, Acceptability, and Appropriateness

All 14 participants completed the psychometric assessment (Table 2). 100% of participants who performed video annotation agreed that the method was feasible, acceptable, and appropriate. Those who performed passive video review reported 67%, 43%, and 50% for feasibility, acceptability, and appropriateness, respectively. Odds ratios favoured the annotation group across all categories: feasibility (4.35, 95% CI: 1.44-13.13, p = 0.009), acceptability (6.04, 95% CI: 1.99-18.33, p = 0.001), and appropriateness (4.01, 95% CI: 1.40-11.52, p = 0.010). Qualitative feedback was also obtained on ways to enhance the learning experience. In the video review group, participants noted a lack of interactivity, limited ability to test oneself, and a need for more focus on key steps. In the annotation group, the addition of anatomical structure identification, incorporating discussion of key decisions, and the ability to highlight errors were components participants believed would enhance the learning experience. Comments also mentioned the annotation of participants’ own operative videos or the opportunity to receive some form of feedback on their annotations would be beneficial. The median time spent on using the material was 120 (IQR: 60-240) minutes and 30 (IQR: 20-120) minutes for the annotation and control groups, respectively (p = 0.12).

**Table 2.** Feasibility, Acceptability and Appropriateness of the Educational Intervention.

| Characteristic | Video annotation | Video review | p- | Difference |
| --- | --- | --- | --- | --- |

|  | group No. (%<br>agreement) | group<br>No. (%<br>agreement) | value | (95% CI) |
| --- | --- | --- | --- | --- |
| **Feasibility** | 28 (100%) | 19 (69%) | 0.001 | 9.00 (4.15-13.84) |
| seems easy | 7 (100%) | 5 (71%) |  |  |
| seems doable | 7 (100%) | 5 (71%) |  |  |
| seems possible | 7 (100%) | 5 (71%) |  |  |
| seems implementable | 7 (100%) | 4 (57%) |  |  |
| **Acceptability** | 28 (100%) | 12 (43%) | 0.000 | 16.00 (10.86-21.13) |
| I welcome this | 7 (100%) | 3 (43%) |  |  |
| I like this | 7 (100%) | 3 (43%) |  |  |
| is appealing to me | 7 (100%) | 3 (43%) |  |  |
| meets my approval | 7 (100%) | 3 (43%) |  |  |
| **Appropriateness** | 28 (100%) | 14 (50%) | 0.000 | 14 (8.82-19.18) |
| seems like a good<br>match | 7 (100%) | 3 (43%) |  |  |
| seems applicable | 7 (100%) | 4 (57%) |  |  |
| seems suitable | 7 (100%) | 3 (43%) |  |  |
| seems fitting | 7 (100%) | 4 (57%) |  |  |

### Outcomes: Knowledge

Step and instrument knowledge scores in the video annotation group improved across all quiz domains, compared to all scores decreasing in the video review group (Table 3). The average change in step knowledge score was higher in the video annotation group compared to the video review group (1, 95% CI: 0.22-1.78, p = 0.017). Overall quiz improvement was also greater in the video annotation group compared to the video review group (1.71, 95% CI: 0.19-3.41, p= 0.048).

When assessing knowledge of sella anatomy, neither group demonstrated significant improvement from baseline. The video annotation group demonstrated a mean improvement in IoU of 1.00% (95% CI: -24.6 to 26.6, p = 0.93), with the video review group obtaining a 9.08% improvement in IoU (95% CI: -14 to 32.86, p = 0.38). No statistically significant difference was observed between the groups (p = 0.39).

### Outcomes: Skills

Both groups demonstrated improvement in mGRS scores from baseline. The video annotation group significantly improved in time and motion (mean difference: 0.79, 95% CI: 0.14–1.43, p = 0.025), instrument handling (mean difference: 0.71, 95% CI: 0.07–1.36, p = 0.035), knowledge of instruments (mean difference: 0.93, 95% CI: 0.37–1.49, p = 0.007), flow of operation (mean difference: 1.43, 95% CI: 0.65–2.20, p = 0.004) and procedural skills knowledge (mean difference: 0.85, 95% CI: 0.22–1.50, p = 0.017). The video review group also imporved across all mGRS subsections, but these imporvements did not reach statistical significance. No significant differences were noted between the groups (Table 4).

**Table 4.**
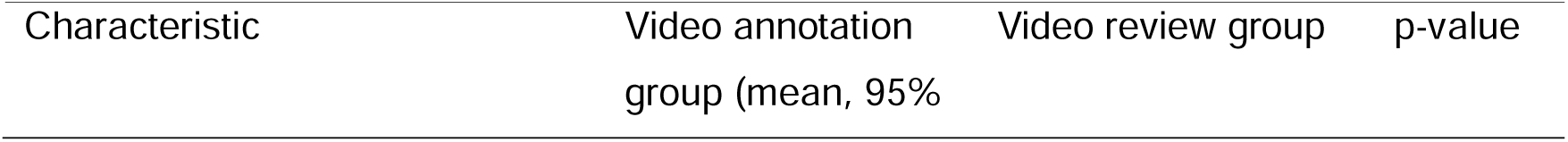
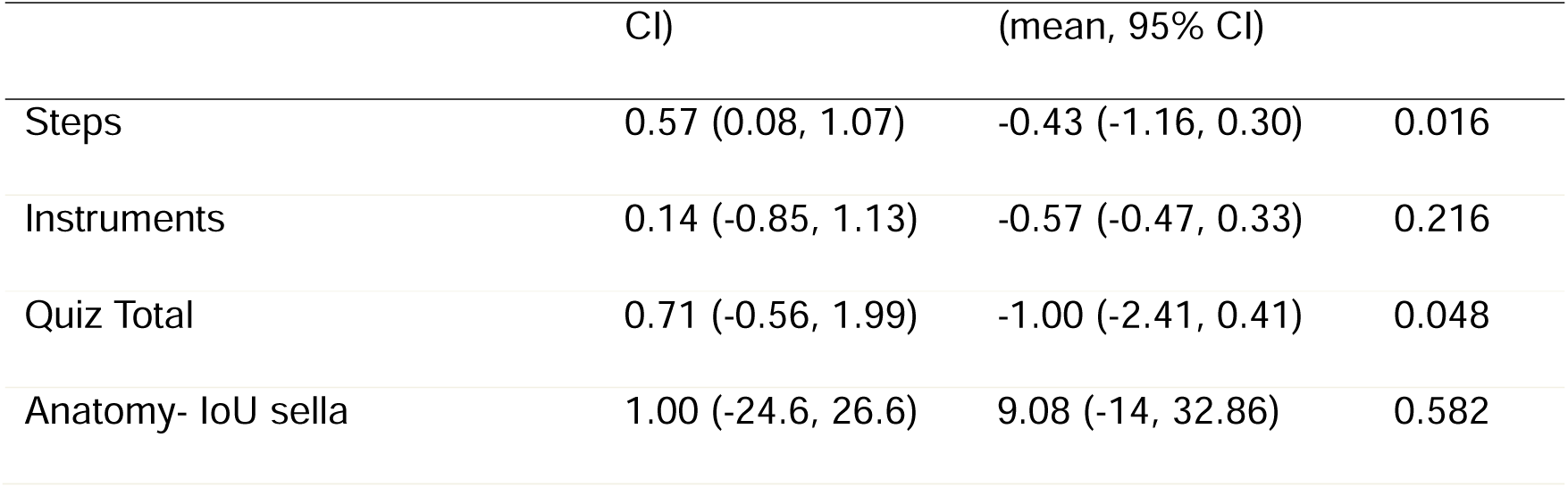
Knowledge Change Between Groups.

| Characteristic | Video annotation<br>group (mean, 95% | Video review group | p-value |
| --- | --- | --- | --- |

|  | CI) | (mean, 95% CI) |  |
| --- | --- | --- | --- |
| Steps | 0.57 (0.08, 1.07) | -0.43 (-1.16, 0.30) | 0.016 |
| Instruments | 0.14 (-0.85, 1.13) | -0.57 (-0.47, 0.33) | 0.216 |
| Quiz Total | 0.71 (-0.56, 1.99) | -1.00 (-2.41, 0.41) | 0.048 |
| Anatomy- IoU sella | 1.00 (-24.6, 26.6) | 9.08 (-14, 32.86) | 0.582 |

### Outcomes: Confidence

From baseline, overall confidence improved significantly in both the video annotation (6 [IQR: 4-9], p = 0.016) and video review groups (6 [IQR: 3-11], p = 0.047) (table 5). Confidence ratings increased in both groups across all domains. Significant improvements were observed in the ability to perform a pituitary adenoma resection category in both the video annotation group (p = 0.016) and the video review group (p = 0.047) and confidence in understanding the decision-making process in the video annotation group (p = 0.031) and the review group (p = 0.031). No significant difference was observed between the groups (table 2).

**Table 5.** Skill Change Between Groups.

| Characteristic | Video annotation group<br>mean, (95% CI) | Video review<br>group | p-value |
| --- | --- | --- | --- |

|  |  | mean, (95% CI) |  |
| --- | --- | --- | --- |
| Respect for Tissue | 0.43 (-0.47-1.33) | 0.14 (-0.55-0.83) | 0.550 |
| Time and Motion | 0.79 (0.14-1.43) | 0.57 (-0.16-1.30) | 0.600 |
| Instrument Handling | 0.71 (0.07-1.36) | 0.28 (-0.41-0.98) | 0.292 |
| Knowledge of Instruments | 0.93 (0.37-1.49) | 0.50 (-0.30-1.30) | 0.305 |
| Flow of operation | 1.43 (0.65-2.20) | 0.64 (-0.35-1.63) | 0.152 |
| Knowledge of Specific Procedure | 0.85 (0.22-1.50) | 0.42 (-0.47-1.33) | 0.3615 |
| Total mGRS | 5.14 (2.36-7.93) | 2.57 (-1.30-6.44) | 0.212 |

**Table 6.** Confidence Change Between Groups.

| Characteristic | Video annotation group median, | Video review group | p-value |
| --- | --- | --- | --- |

|  | (IQR) | median, (IQR)) |  |
| --- | --- | --- | --- |
| I feel confident in being able to perform a pituitary adenoma resection | 2 (2-3) | 2 (1-3) | 0.715 |
| I feel confident in identifying key anatomical structures involved in this procedure | 1 (0-2) | 1(0-3) | 0.639 |
| I feel confident in understanding the key technical steps required to perform this procedure. | 1 (1-2) | 1 (0-3) | 0.864 |
| I feel confident in understanding the decision-making process behind the key technical steps involved in this procedure. | 1 (1-2) | 1 (1-2) | 0.849 |
| I feel confident highlighting the common potential errors that can occur while undertaking this procedure. | 1 (0-2) | 1 (0-2) | 0.904 |
| Total | 6 (4-9) | 6 (3-11) | 0.899 |

## Discussion

### Principal Findings

This study aimed to assess the impact of active operative video annotation on surgical learning, exploring the domains of implementability, knowledge, skills, and confidence. Video annotation as a learning tool demonstrated highly favourable implementability, with strong agreement for feasibility, acceptability, and appropriateness, when compared to passive video review. The video annotation group also showed favourable step and overall procedural knowledge scores compared to the passive video review group. Significant improvements in surgical skills were observed in the video annotation group, which were not seen in the video review group, although no significant differences between the groups were noted. Both groups showed improved confidence, but again, no significant difference between the groups. This study is the first to assess the impact of video annotation of steps and instruments on learning outcomes potentially offering a widely accessible, cost-effective method for surgeons and trainees to adapt to the changing surgical training landscape.

### Findings in the Context of the Literature

The use of operative video in surgical training has been shown to increase attention. In particular, operative videos are often chosen in preference to non-operative videos on free video-sharing platforms when engaging in self-directed learning (24). A variety of methods for implementing operative surgical videos exist (13, 25), including their unstructured narration (26), expert step editing (27), the application of cognitive task analyses (21), and interactive viewing methods (28). Despite its widespread use in the literature, few studies assess such interventions across all domains of confidence, knowledge, skills, and implementability (13). Surgical procedures, especially those involving minimally invasive techniques, endoscopy, or robotics, have a steep learning curve. In the current working environment, characterised by time restrictions, the impact of COVID-19, and increasing expectations, there is now a significant training void. Video annotation could help address this gap, ensuring patient safety and trainee competency.

This study is the first application of a validated psychometric assessment tool to evaluate the implementability of a surgical video intervention for education. Our findings align with existing literature, showing that operative video interventions are generally reported as easy to use, are good educational tools, and enjoyable (29). Two studies on video use in surgical education (30, 31) used a validated tool, the Heidelberg Inventory for the Evaluation of Teaching Courses (HITEC) tool, which primarily focuses on evaluating instructors and didactic courses. They found favourable results for the multimedia and video based intervention (30) although with no significant difference compared to printed format (31). In our study, video annotation did not significantly increase learning time, a crucial consideration for novice surgeons managing demanding schedules. Previous research supports the idea that interactive teaching methods reduce preparation time by enhancing motivation and engagement (32) (33). The 100% agreement regarding feasibility and ease of use also suggests that this platform is user-friendly for novice surgeons. Although there are challenges in implementing new and complex technologies such as virtual reality, our findings suggest that video annotation is an accessible alternative, particularly in resource-limited settings (29, 34).

Interactive learning methods have demonstrated effectiveness in increasing surgical knowledge retention (35) (36) (32), consistent with our findings of improved procedural knowledge. However, the precise anatomy of the sella remains challenging, even for expert neurosurgeons (19), which may explain the limited changes observed in anatomical knowledge. The video annotation exercise also did not include an anatomy labelling component. Surgical skill development involves the incorporation of a number of different learning methods, of which learning through observation plays a crucial role (37). Our study found improvements from baseline in the video annotation group, likely attributable to both the active action of video annotation and practice on simulators. This suggests that the combination of interactive cognitive training and motor skill practice may enhance learning outcomes more than passive video watching and simulation practice alone, which did not produce the same level of improvement. The study by Cizmic, Häberle (38) also supports the idea that multiple iterations of operative video annotation might continue to impact surgical learning and improve the learning curve over time. Confidence is a subjective measure, and mixed findings have been reported when assessing its correlation with surgical skill performance (39, 40). In our study, both groups demonstrated similar improvements confidence, although it is important to consider that observing videos can create an illusion of learning (41).

### Strengths and Limitations

This is the first study, to the knowledge of the author, that evaluates the use of step and instrument video annotation for surgical training. This study comprehensively evaluated implementability outcomes, knowledge, skills, and confidence, a combination which is rarely seen in previous studies (13). The tools used for evaluating surgical learning outcomes were appropriate. The knowledge test utilised real-life operative videos and included multiple images for annotation and multiple videos for step identification to improve reliability. The mGRS is well-validated and provides an objective assessment of participant performance. The blinding of assessors and participants, along with the use of two markers to evaluate simulation performance, significantly reduced bias. By comparing video annotation to video review, we were able to isolate the impact of video annotation on learning outcomes while accounting for the natural progression of surgical skills through repetitive simulations. The study’s simulated environment may limit its generalizability to real-world surgical performance (42) or patient outcomes (43). The video annotation method did not incorporate annotation of sellar anatomy, which may have limited learning in this area. Additionally, the study was underpowered to detect the effect sizes observed. Multiple variables were tested for improved learning outcomes which may increase the risk of type 1 error. Future studies should focus on the impact of video annotation as a learning method over time to assess its effect on the learning curve, particularly in both senior and more junior cohorts.

### Conclusion

We examined the implementability of active video annotation and its impact on surgical learning across the domains of knowledge, skills, and confidence. Video annotation was rated very highly for feasibility, acceptability, and appropriateness. Passive video review was also generally considered to be implementable. Active video annotation led to a higher change in knowledge scores compared to passive video review and also improved skills and confidence from baseline. Its impact, however, on real-world surgical performance and patient outcomes requires further study.

## Data Availability

All data relevant to the study are included in the article.

## Acknowledgements/funding

Mr Marcus is funded by the NIHR Biomedical Research Centre at University College London. Mr Khan is supported by the NIHR Academic Clinical Fellowship

## Data statement

All data relevant to the study are included in the article.

